# The impact of pausing the Oxford-AstraZeneca COVID-19 vaccine on uptake in Europe: a difference-in-differences analysis

**DOI:** 10.1101/2021.08.30.21262821

**Authors:** Vageesh Jain, Paula Lorgelly

**Affiliations:** Institute for Global Health, University College London, London, WC1E 6JB, UK; Institute of Epidemiology and Health Care, University College London, London WC1E 7HB, UK

**Author notes:** Corresponding author: Vageesh Jain, Address: Institute for Global Health, University College London, London, WC1E 6JB.

**Keywords:** Public health, COVID-19, health policy

## Abstract

**Background:** Several countries paused their rollouts of the Oxford-AstraZeneca COVID-19 vaccine in mid-March 2021 due to concerns about vaccine-induced thrombosis and thrombocytopenia. Many warned that this risked damaging public confidence during a critical period of pandemic response. This study investigated whether the pause in the use of the Oxford-AstraZeneca vaccine had an impact on subsequent vaccine uptake in European countries.

**Methods:** We used a difference-in-differences approach capitalizing on the fact that some countries halted their rollouts whilst others did not. A longitudinal panel was constructed for European Economic Area countries spanning 15 weeks in early 2021. Media reports were used to identify countries that paused the Oxford-AstraZeneca vaccine and the timing of this. Data on vaccine uptake were available through the European Centre for Disease Control and Prevention COVID-19 Vaccine Tracker. Difference-in-differences linear regression models controlled for key confounders that could influence vaccine uptake, and country and week fixed effects. Further models and robustness checks were performed.

**Results:** The panel included 28 countries, with 19 in the intervention group and 9 in the control group. Pausing the Oxford-AstraZeneca vaccine was associated with a 0.52% decrease in uptake for the first dose of a COVID-19 vaccine and a 1.49% decrease in the uptake for both doses, comparing countries that paused to those that did not. These estimates are not statistically significant (p=0.86 and 0.39 respectively). For the Oxford-AstraZeneca vaccine only, the pause was associated with a 0.56% increase in uptake for the first dose and a 0.07% decrease in uptake for both doses. These estimates are also not statistically significant (p= 0.56 and 0.51 respectively). All our findings are robust to sensitivity analyses.

**Conclusion:** As new COVID-19 vaccines emerge, regulators should be cautious to deviate from usual protocols if further investigation on clinical or epidemiological grounds is warranted.

## Introduction

A number of European countries paused their national rollouts of the Oxford-AstraZeneca COVID-19 vaccine in mid-March 2021(1–3), due to concerns about a rare but serious syndrome known as vaccine-induced thrombosis and thrombocytopenia (VITT). According to the European Medicines Agency (EMA), 30 cases of thromboembolic events (predominantly venous) had been reported by March 10, 2021, among the approximately 5 million recipients of the Oxford– AstraZeneca COVID-19 vaccine in the European Economic Area (EEA). The EMA subsequently stated, “The number of thromboembolic events in vaccinated people is no higher than the number seen in the general population”(4). The World Health Organization (WHO) also recommended continuing vaccination whilst further data was awaited and reviewed(5). Given the uncertainty around a causal relationship, many criticised regulatory intervention to halt vaccination as unnecessarily damaging to public confidence, during a critical period of epidemic response(6). A YouGov poll covering around 8,000 people in seven European countries from March 12^th^ to 18^th^ found that in France, Germany, Spain and Italy, people were more likely to see the AstraZeneca vaccine as unsafe than as safe(7), unlike in previous surveys(8).

After a short pause of approximately one week, most EEA nations resumed their use of the vaccine(9,10), although the Scandinavian countries did not. Sweden, Finland and Iceland moved toward limiting the use of the Oxford-AstraZeneca vaccine for their elderly population, while Denmark and Norway prolonged their suspension of the AstraZeneca vaccine and shifted to providing alternative vaccines(11). This study seeks to understand whether the pause in the use of the Oxford-AstraZeneca vaccine had a significant impact on uptake for both the Oxford-AstraZeneca vaccine specifically, or on COVID-19 vaccine uptake more broadly. We were able to use quasi-experimental difference-in-differences methods as the vaccination pause occurred in some European countries but not others, at approximately the same time and for a similar duration across countries. With another generation of COVID-19 vaccines on the horizon, this study will help to inform future regulatory processes in the context of novel technologies used in pandemic response.

## Methods

### Data Sources

A longitudinal panel was constructed for 28 EEA countries with vaccine uptake data at three-week intervals from week 3 to 18 of the COVID-19 vaccination rollout in 2021. This did not include the United Kingdom (UK), Denmark or Norway. The UK is no longer in the EEA and had different plans and systems for vaccine rollout compared to the rest of the EEA. Denmark and Norway did not restart their use of the Oxford-AstraZeneca vaccine after the pause, unlike other countries. Media reports were used to identify countries that paused the Oxford-AstraZeneca vaccine and the timing of this(1–3). Data on overall COVID-19 vaccine uptake and only Oxford-AstraZeneca uptake, for first and both doses, were obtained directly from the European Centre for Disease Control and Prevention COVID-19 Vaccine Tracker and Open Data(12,13). Uptake was provided as a cumulative percentage of the adult population (18+) for all vaccines, and due to data limitations, calculated as a cumulative percentage of the entire population for the Oxford-AstraZeneca vaccine.

Data on numbers of vaccines supplied, for both the Oxford-AstraZeneca vaccine alone and for all vaccines, were also obtained from the same dataset and adjusted for the population size of each country, prior to analysis. Data on other potential confounders, including demographic, economic, and health system indicators, were from 2019 and collected from the World Bank Open Database(14). Two indicators (universal health coverage service index and health expenditure per capita) were from 2017 and 2018 respectively. Given the role of public confidence and trust in vaccine uptake, the state of democracy in included countries was ascertained through the Economist Intelligence Unit’s 2020 Democracy Index(15). Data on weekly COVID-19 cases and cumulative deaths at baseline (week 3 of 2021) were obtained from the open-access Our World in Data(16).

### Exposures and Outcomes

Exposure to the intervention was defined as those EEA countries that paused the rollout of the Oxford-AstraZeneca vaccine. This action was taken over the week of 8^th^ March. Those that did not pause the vaccine at all were treated as the control group. A dummy variable was generated to indicate whether the vaccine rollout was paused in a given country at a given week of the vaccination programme. Our primary outcome was cumulative uptake for first and both doses for all vaccines, defined as the percentage of the adult (18+) population that had received 1) one dose and 2) both doses of a COVID-19 vaccine, from week 3 of 2021 to week 18. The secondary outcome was cumulative uptake for first and both doses of the Oxford-AstraZeneca vaccine from week 6 to 18. Due to data availability this was defined as the percentage of the whole population that had received 1) one dose and 2) both doses of the Oxford-AstraZeneca vaccine.

### Statistical Analysis

All analyses were conducted using STATA 14(17). We first compared both groups of countries on a range of baseline characteristics, using the Wilxocon Rank Sum test to identify statistically significant differences in mean values between groups. P-values of less than 0.05 were considered statistically significant. We then fit multivariate linear regression models in which the key variable of interest was a difference-in-differences estimator, interpreted as the average effect of pausing the Oxford-AstraZeneca vaccine on overall vaccine uptake, in the countries that paused it. Our main model that investigated uptake of all vaccines as the primary outcome controlled for weekly supplies of all vaccines per population, and the model investigating the impact on uptake of the Oxford-AstraZeneca vaccine only controlled for weekly supplies per population of this specific vaccine. This was deemed to be a critical and observed time-varying confounder, which also varied across countries despite joint procurement plans, therefore being potentially correlated with differences in vaccine uptake between the countries that paused the vaccine rollout and those that did not. We included both country fixed effects to control for underlying time-invariant differences between countries, and week fixed effects to account for changes over time common to all countries.

We ran several further checks to assess whether our results were robust and to probe the assumptions that support the methodology, including the parallel trends assumption, which states that in the absence of the pause in vaccine rollout, included countries would have had the same trends in vaccine uptake as control countries. We first constructed graphs to examine the trends in average vaccine uptake before the pause. We then ran a statistical test for parallel trends using the ‘estat ptrends’ command in STATA 17(18). We ran a second model also adjusting for country and week fixed effects, to exclude countries that imposed later age restrictions on the use of the Oxford AstraZeneca vaccine from early April(19–23): Belgium, Estonia, Finland, France, Germany Iceland, Ireland, Italy, the Netherlands, Portugal, Sweden and Spain. We also ran a third multivariable model using the random-effects estimator and adding time-invariant covariates that were statistically significantly different between both groups of countries on prior hypothesis testing, including GDP per capita and EIU democracy index. In all models, similar results were obtained (Supplementary File).

Data was collected at the country-level and all countries were part of the EEA trading bloc, part of a joint vaccine procurement and distribution strategy(24). The difference-in-differences panel estimator could therefore be subject to serial autocorrelation, within-cluster correlation and cross-sectional dependence, potentially leading to standard errors that are not robust. We used clustered standard errors in all models to account for this.

## Results

A total of 30 EEA countries were initially selected but two were excluded, leaving 28 for the primary analysis. The intervention group consisted of 19 countries and the remaining 9 countries that did not pause the vaccine rollout formed the control group. A list of countries in each group is provided in the Supplementary File.

Linear trends are parallel in both groups, with no evidence against the null hypothesis (p=0.56 for overall uptake of first dose, p=0.67 for overall uptake of both doses; p=0.53 for Oxford-AstraZeneca first dose and p=0.79 for Oxford-AstraZeneca both doses). The general trend for vaccination uptake across all countries from week 3 to 18 of 2021 was upward, although the gradient of the increase was steeper when looking at all vaccines, compared to Oxford-AstraZeneca alone, and for first doses compared to both doses (Figure 1).

**Figure 1.**
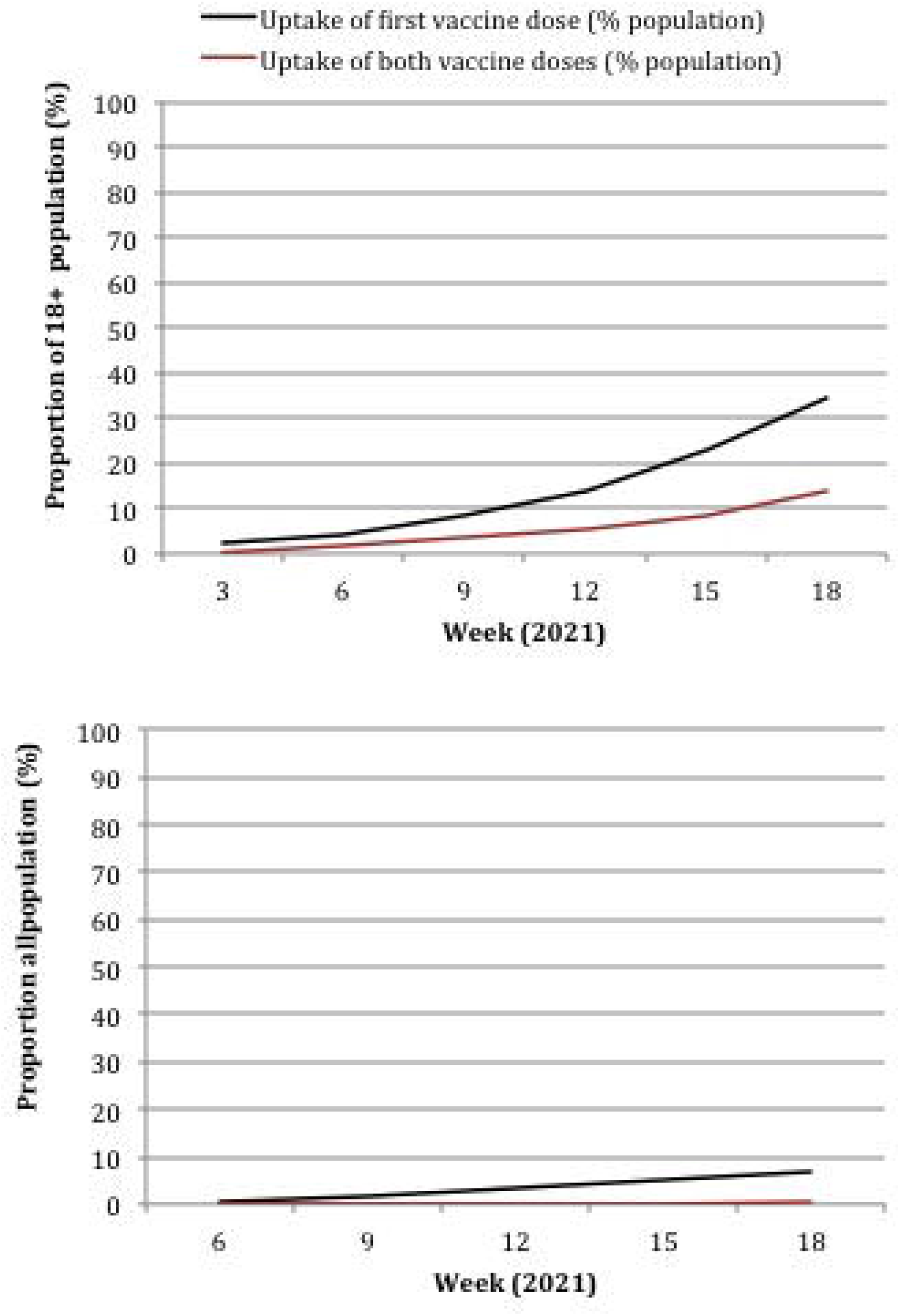
All vaccines (top) and Oxford-AstraZeneca vaccine (bottom) uptake over time across all included EEA countries.

Table 1 shows the demographic, socioeconomic, and health system characteristics, as well as data on COVID-19 epidemics at baseline, presented as means (95% CI), for each group of countries. On average, countries that paused the vaccine had higher GDP per capita and stronger democratic performance, compared to those that did not. By controlling for these time-invariant covariates we adjusted for these differences between countries in the difference-in-differences analysis. Population size, age, gender, life expectancy at birth and COVID-19 epidemic characteristics at baseline (measured as a 7-day average COVID-19 case rate and cumulative death rate per million in the week up to 21^st^ January) were not significantly different between both groups of countries.

**Table 1.** Baseline characteristics between intervention and control groups

| Domain | Indicator | Control (n=9) | Intervention (n=19) | P-value (difference) |
| --- | --- | --- | --- | --- |
| Demographic | Population (millions) | 10.3 (1.58 – 18.9) | 18.4 (6.01 – 30.8) | 0.90 |
|  | Age>=65 (% total population)* | 20.3 (19.2 – 21.3) | 18.9 (17.6 – 20.3) | 0.31 |
|  | Age 0-14 (% total population)* | 15.0 (14.1 – 15.9) | 16.1 (15.0 – 17.1) | 0.17 |
|  | Male (% total population)* | 48.8 (48.2 – 49.3) | 48.8 (48.2 – 49.5) | 0.45 |
| Socioeconomic | Urban population (% total population) | 68.3 (50.2 – 88.4) | 72.8 (66.4 – 79.2) | 0.82 |
|  | Unemployment (% total labour force) | 5.29 (1.64 – 8.93) | 6.08 (4.82 – 7.35) | 0.06 |
| | GDP per capita (current International \$, PPP)* | 37274.1 (29062.3 – 45486.9) | 51430.7 (41085.1 – 61776.3) | 0.04 |
| Political | EIU Democracy Index* | 7.11 (6.69 – 7.53) | 8.09 (7.68 – 8.51) | 0.006 |
| Health | Current health expenditure per capita (PPP, current International \$)* | 2790.4 (1716.2 – 3864.6) | 4043.4 (3263.3 – 4823.4) | 0.05 |
|  | Universal health coverage service index* | 75.4 (70.6 – 80.1) | 79.3 (77.2 – 81.4) | 0.08 |
|  | Life expectancy at birth (total years) | 79.6 (77.2 – 81.9) | 80.5 (79.2 – 81.9) | 0.55 |
| COVID-19 Epidemic | 7-Day average COVID-19 cases per million at baseline (week 3) | 213.5 (55.6 – 367.4) | 350.4 (226.7 – 474.0) | 0.09 |
|  | Cumulative COVID-19 deaths per million at baseline (week 3) | 1126.9 (817.9 – 1435.8) | 773.5 (574.0 – 973.0) | 0.06 |

\*No data available for Liechtenstein

Figures 2 and 3 show line graphs for first and both dose vaccine uptake over time comparing countries that paused with those that did not. For overall COVID-19 vaccination the trend in both groups is similar for the first dose, with overlapping confidence intervals at all time points and no deviation after the pause. For both doses there appears to be some deviation after the pause, with countries that paused having marginally lower rates of uptake, but confidence intervals suggested that these differences were non-significant. For the Oxford-AstraZeneca vaccine, uptake was similar in both groups over time with minimal deviation after the pause. Although the population denominators for vaccine uptake were not directly comparable, the Oxford-AstraZeneca vaccine contributed little to overall vaccine uptake. This was particularly pronounced when looking at both doses, suggesting there was a temporal shift away from using the Oxford-AstraZeneca vaccines in all included EEA countries.

**Figure 2.**
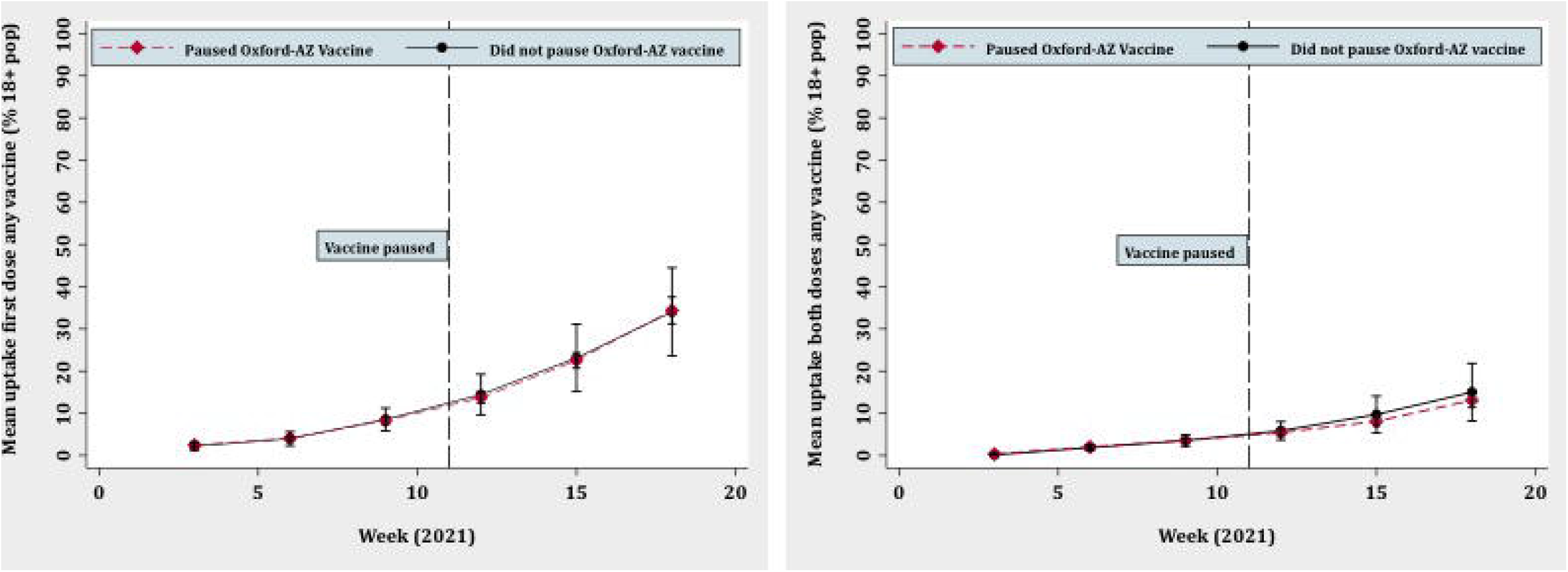
Overall COVID-19 vaccine uptake over time for first dose (left) and both doses (right): countries that paused Oxford-AstraZeneca vaccine vs. those that did not.

**Figure 3.**
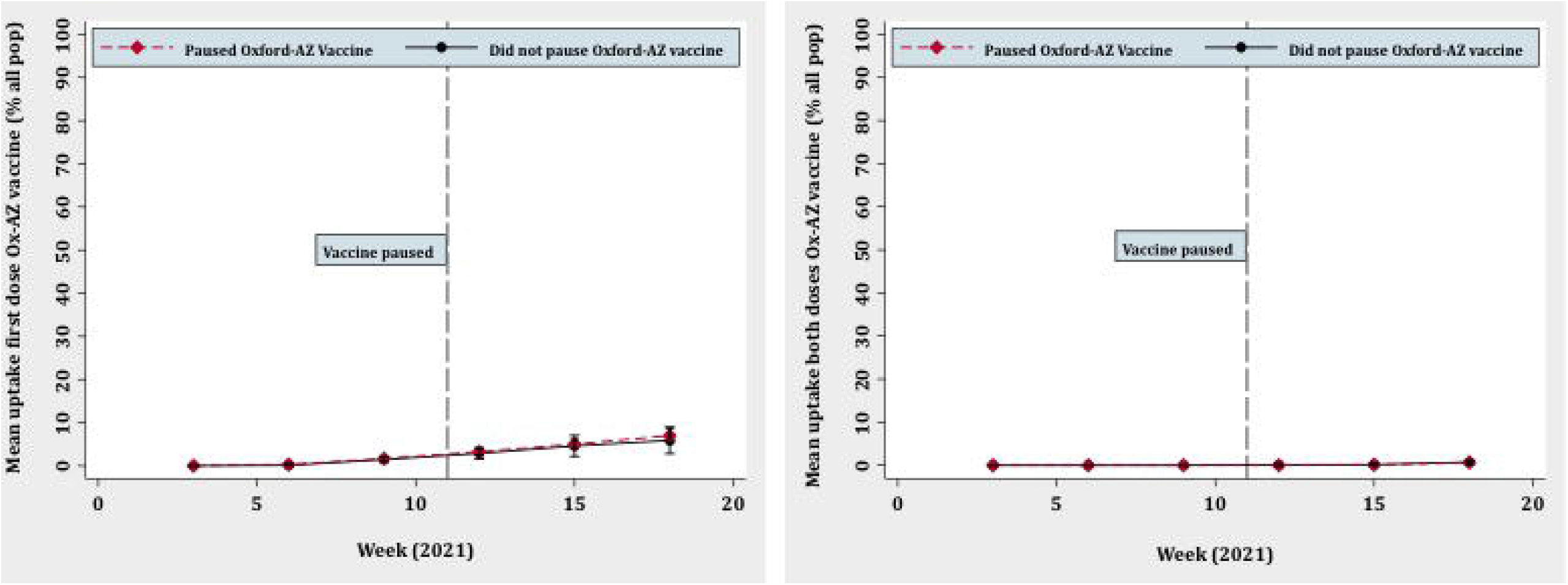
Oxford-AstraZeneca vaccine uptake over time for first dose (left) and both doses (right): countries that paused Oxford-AstraZeneca vaccine vs. those that did not.

Our difference-in-differences estimates found that pausing the rollout of the Oxford-AstraZeneca vaccine was associated with a subsequent 0.52% decrease in the overall population uptake for the first dose of a COVID-19 vaccine and a 1.49% decrease in the uptake for both doses, comparing countries that paused to those that did not. These estimates are not statistically significant (p=0.86 and 0.39 respectively). When looking at the uptake of the Oxford-AstraZeneca vaccine only, the pause was associated with a 0.56% increase in uptake for the first dose and a 0.07% decrease in uptake for both doses. These estimates are also not statistically significant (p= 0.56 and 0.51 respectively). These findings are robust to sensitivity analyses including the exclusion of some countries and the use of the random-effects estimator (Supplementary File) suggesting that the pause had no significant impact on subsequent vaccine uptake.

## Discussion

### Key Findings

There was no significant difference in subsequent vaccine uptake in countries that paused the Oxford-AstraZeneca vaccine in mid-March compared with those that did not. This was true for both one and two doses of the vaccine, and when looking at overall vaccine uptake (including all vaccines) and the Oxford-AstraZeneca vaccine alone. This suggests that over time, public confidence in vaccination was not significantly more damaged in those countries that paused the vaccine compared to those that did not. It appears there has been a shift away from using the Oxford-AstraZeneca vaccine in the EEA due to relatively low rates of uptake as a proportion of all vaccine uptake and a shallower increasing gradient in uptake over time compared to all vaccines, but this trend was similar across all included countries. This analysis suggests that as new COVID-19 vaccines emerge, regulators should be cautious to deviate from usual pharmacovigilance protocols on the sole basis of potential impacts on public confidence, if further investigation on clinical or epidemiological grounds is warranted.

### Explaining findings

Many experts were concerned about the impact of pausing the Oxford-AstraZeneca vaccine on public confidence and consequent demand(6). Regulators that paused the rollout highlighted that VITT was serious but rare. Media and scientific communications emphasised the fact that investigating safety signals in this manner is an established phamacovigilance process done out of an abundance of caution, reiterating the well-established safety and effectiveness of vaccine(25). For instance, after pausing the vaccine Spain’s health minister Carolina Dias said “We take this decision today in the interest of caution,” referring to “infrequent cases which are very few but very significant and have prompted Spain to join the other countries that have opted for this precautionary suspension” (26). Around the same time, the UK Medicines and Healthcare products Regulatory Agency (MHRA) reported that VITT was seen in less than 1 per million vaccinated people in the UK, which had administered more Oxford-AstraZeneca doses than other European countries, and that a causal association had not been established(27). Most countries restarted using that vaccine approximately one week later, serving to further reassure the public in countries that initially paused the vaccine. It is likely that a short pause, variation in regulatory practice across countries, and reassuring public communications all helped to mitigate any potential impact on vaccine uptake in countries that did pause their rollouts.

Vaccine uptake may also have been similar across both groups of countries because of later scientific and policy developments on the Oxford-AstraZeneca vaccine. Although the majority of countries that had initially paused the vaccine in mid-March restarted it shortly after, 12 out of 28 included countries restricted its use in younger people from the second week of April(20–22,28). This was in response to concern about the balance of risks and benefits in the young. In these countries, the later impact of the pause on vaccine uptake (at weeks 15 and 18 in our study) may have been blurred by the impact of these restrictions. We did however control for vaccine supply over time across countries, which can be considered a proxy for the impact of such additional regulations since countries would be unlikely to procure vaccines they were not planning to use. A sensitivity analysis (Supplementary File) excluding the countries that imposed additional later restrictions on the use of the Oxford-AstraZeneca vaccine also showed that the results did not change. The relatively timid increase over time in uptake for both doses of the Oxford-AstraZeneca vaccine is likely a product of timing (with the second dose administered at 4-12 weeks after the first(29)) as well as these additional restrictions. Importantly, the same trend was seen in both countries that paused the vaccine and those that didn’t, suggesting that countries that paused the vaccine were not at a later disadvantage compared to others.

### Strengths and Limitations

We were able to answer this critical public health policy question using a natural experiment, requiring no manipulation to allocate the intervention. All included countries were part of the EEA and therefore to some extent comparable, and our statistical analyses accounted for important differences across groups of countries where present. The intervention itself occurred at approximately the same time across countries, with a direct and immediate impact on both public information and national vaccine programme rollouts, further supporting our approach. Outcome data were available from a single and reliable source (the ECDC) meaning our dataset was largely complete and of a high quality. Through investigating the impact on uptake for all vaccines as well as the Oxford-AstraZeneca vaccine specifically, our study was well designed to understand the various mechanisms through which the pause could have affected vaccine uptake.

Our study had several limitations. First, although we were able to allocate countries to one of two groups, it is possible that there was contamination from the intervention to the control group. The European medicines regulatory system is based on a network of around 50 regulatory authorities from across EEA countries(30). Citizens may place more trust in their own national regulators compared with others. But populations in countries that did not pause the Oxford-AstraZeneca vaccine may have also been affected by knowing that similar countries (operating under a common European regulatory framework) decided to pause their rollouts of the vaccine, despite it being restarted soon after. While this could theoretically minimise any observed differences in vaccine uptake between countries, this is an important corollary of the intervention in a complex global regulatory environment during a pandemic, which should not be artificially adjusted for but considered part of the intervention itself. Second, although we were able to include data spanning 15 weeks, we were unable to assess the long-term impact of the pause on vaccine uptake, although for this particular question a short timeframe was deemed most appropriate to assess impact. Third, the difference-in-differences methodology relies on the assumption that there are no factors beyond the pausing of the Oxford-AstraZeneca vaccine that differentially affect vaccine uptake and thereby confound the observed effect. We accounted for these through adjusting for fixed country and week effects, adding time-variant fluctuations in supplies of vaccines across countries. We also controlled for other potential time-invariant confounders through a random-effects model. Nevertheless, the omission of other important, unobserved, time-varying characteristics could bias our findings if they affected the intervention and control group differentially. Finally, although difference-in-differences is an often used quasi-experimental method(31), inferences about causality can be less definitive than those obtained through a well designed randomised controlled trial. But it would not be possible to evaluate the impact of pausing the rollout of a vaccine during a pandemic, making quasi-experimental methods such as those used here the gold standard for policy evaluation.

### Implications for research and policy

Although we found small non-significant decreases in overall vaccine uptake in countries that paused the Oxford-AstraZeneca vaccine compared to those that did not, we found a small non-significant increase in uptake for the first dose of the Oxford-AstraZeneca vaccine itself. Further research is required to understand whether populations feel more reassured after they know regulators have closely scrutinised the data on a particular vaccine and whether this paradoxically puts it at an advantage compared to others in terms of public perception and attitudes.

Despite arguments made against pausing the Oxford-AstraZeneca vaccine rollout, we found no evidence that it significantly impacted vaccine uptake. The findings from this study cannot however be generalized to future scenarios where vaccine risks, the stage of the pandemic, the mechanism and duration of pausing rollout, public perception of the regulators that pause and those that do not, and public communications could all be different. Future qualitative research will be needed to understand how these factors affect the impact of a nationwide pause in the rollout of a novel vaccine, and which policies and plans can be put in place to pause vaccines in as safe a way as possible, allowing further investigation whilst preserving public confidence. This analysis also highlights the general lack of alignment between European decision-making bodies, despite the fact that the public health impact of a regionally uncoordinated approach to stopping, restarting, and later restricting vaccine use, was largely unknown. Policymakers and regulatory authorities in Europe must collaborate through regional organizations like the EMA and global organizations like the WHO, to ensure that in the context of an international health threat, national policies and processes on vaccines are harmonised as far as possible. In the long-term this will be necessary to promote public trust in COVID-19 vaccines and in the institutional processes related to their selection, procurement, distribution and monitoring.

## Conclusions

In this quasi-experimental study we employed a difference-in-differences design to estimate the causal effect of pausing the Oxford-AstraZeneca vaccine on later vaccine uptake. We found that there was no significant difference in overall COVID-19 vaccine uptake in countries that paused the Oxford-AstraZeneca vaccine in mid-March compared with those that did not. There was a shift away from using the Oxford-AstraZeneca vaccine itself, but uptake was not significantly different across countries. Further research is required to understand the precise mechanism underlying our findings, and the factors associated with how the public within and across countries perceive a pause in the national rollout of a novel vaccine. As new COVID-19 vaccines emerge, regulators should be cautious to deviate from usual pharmacovigilance protocols if further investigation on clinical or epidemiological grounds is warranted.

## Data Availability

Data underpinning this analysis will be available as a supplement once the paper has been peer-reviewed.

## Authors and Contributors

Substantial contributions to the conception or design of the work; or the acquisition, analysis, or interpretation of data for the work: VJ, PL

Drafting the work or revising it critically for important intellectual content: VJ, PL

Final approval of the version to be published: VJ, PL

Agreement to be accountable for all aspects of the work in ensuring that questions related to the accuracy or integrity of any part of the work are appropriately investigated and resolved: VJ, PL

VJ conceptualized and designed the study and completed data collection and curation. The methodology was discussed and agreed between VJ and PL. Data analysis and interpretation was first done by VJ and then validated independently by PL through to confirm the validity of the approach and findings. Project administration, data visualisation and writing of the original draft was done by VJ and review and editing by PL who provided overall supervision for the project.

