## Supplementary File for "The impact of pausing the Oxford-AstraZeneca COVID-19 vaccine on uptake in Europe: a difference-in-differences analysis"

**Table 1 - Country groupings**

| Control Group | Intervention Group |
| --- | --- |
| Belgium | Austria |
| Bulgaria | Cyprus |
| Croatia | Estonia |
| Czech Republic | Finland |
| Greece | France |
| Hungary | Germany |
| Liechtenstein | Iceland |
| Malta | Ireland |
| Poland | Italy |
|  | Latvia |
|  | Lithuania |
|  | Luxembourg |
|  | Netherlands |
|  | Portugal |
|  | Romania |
|  | Slovakia |
|  | Slovenia |
|  | Spain |
|  | Sweden |

**Regression models and sensitivity analyses**

**Table 2 – Overall COVID-19 vaccine uptake**

| Model | Country-level covariates | Fixed/random effects | Change in overall COVID-19 vaccine uptake for first dose (%) | Robust Standard Error | P-Value | Change in overall COVID-19 vaccine uptake for both doses (%) | Robust Standard Error | P-Value | Number of observations |
| --- | --- | --- | --- | --- | --- | --- | --- | --- | --- |
| 1 (Main) | Vaccine supply | Country and week fixed-effects | -0.52 | 2.82 | 0.86 | -1.49 | 1.72 | 0.39 | 167 |
| 2* | Vaccine supply | Country and week fixed-effects | -2.09 | 3.30 | 0.54 | -1.74 | 2.00 | 0.40 | 95 |
| 3 | Vaccine supply, GDP per capita, EIU Democracy Index | Random effects | -0.70 | 3.10 | 0.82 | -1.47 | 1.93 | 0.45 | 162 |

* Included countries: Austria, Bulgaria, Croatia, Cyprus, Czech Republic, Greece, Hungary, Latvia, Liechtenstein, Lithuania, Luxembourg, Malta, Poland, Romania, Slovakia, Slovenia

**Table 3 – Oxford-AstraZeneca vaccine uptake**

| Model | Fixed/random effects | Fixed/random effects | Change in overall COVID-19 vaccine uptake for first dose (%) | Robust Standard Error | P-Value | Change in overall COVID-19 vaccine uptake for both doses (%) | Robust Standard Error | P-Value | Number of observations |
| --- | --- | --- | --- | --- | --- | --- | --- | --- | --- |
| 1 (Main) | Vaccine supply | Country and week fixed-effects | 0.56 | 1.02 | 0.59 | -0.07 | 0.13 | 0.59 | 168 |
| 2* | Vaccine supply | Country and week fixed-effects | 0.65 | 1.03 | 0.54 | -0.13 | 0.12 | 0.31 | 96 |
| 3 | Vaccine supply, GDP per capita, EIU Democracy Index | Random effects | 0.02 | 0.99 | 0.98 | -0.12 | 0.13 | 0.35 | 162 |

* Included countries: Austria, Bulgaria, Croatia, Cyprus, Czech Republic, Greece, Hungary, Latvia, Liechtenstein, Lithuania, Luxembourg, Malta, Poland, Romania, Slovakia, Slovenia
